# The misleading illusion of COVID-19 confirmed case data: alternative estimates and a monitoring tool

**DOI:** 10.1101/2020.05.20.20107516

**Authors:** Rogelio Macías-Ordóñez, Damián Villaseñor-Amador

## Abstract

*Confirmed Case Data* have been widely cited during the current COVID-19 pandemic as an estimate of the spread of the SARS-CoV-2 virus. However, their central role in media, official reports and decision-making may be undeserved and misleading. Previously published Infection Fatality Rates were weighted by age structure in the 50 countries with more reported deaths to obtain country-specific rates. For each country, the number of infections up to the Infection Date (23 days ago = Incubation Period + Onset to Death period) and the present percentage of immune population were estimated using Infection Fatality Rate, the number of reported deaths (which is less prone to undersampling), and projecting back to Infection Date. We then estimated a Detection Index for each country as the percentage of estimated infections that confirmed cases represent. Assuming that detection remains constant after Infection Date, we estimated the number of deaths and the estimated percentage of the population of each country expected to be immune up to 23 days into the future. Estimated Infection Fatality Rates are higher in Europe. In most countries, confirmed cases currently represent less than 30% of estimated infections on Infection Date, and this value decreases with time. Countries with flat curves throughout the pandemic show the lowest immunity percentages and these values seem unlikely to change in the near future, suggesting that they remain vulnerable to new outbreaks. Estimates for some countries with low Infection Fatality Rates suggest a still steep increase in the number of casualties in the next three weeks. Countries that did not control initial outbreaks seem to have reached higher immunity percentages, although mostly still under 5%. We provide the code to monitor the trajectories of these estimates in 178 countries throughout the COVID-19 pandemic.

## Introduction

COVID-19 *confirmed case data* (CCD) are the central piece of information in most news, official reports, conversations, forecasting efforts, and are also probably central to most decisions made by authorities worldwide since the pandemic outbreak in December 2019. However, it is widely known that they represent a small and unknown fraction of the actual number of SARS-CoV-2 infections [1 -4], we just do not know how small. Especially hard to assess is the number of asymptomatic but contagious infections [5-8], as asymptomatic carriers are unlikely to seek testing.

Elementary sampling theory and recent supported opinions [3,6,9] suggest that CCD are highly dependent on the testing effort and sampling protocol, among other factors. Unless a randomized and standardized sampling protocol of the whole population is carried out, there is no *a priori* reason to assume they are representative of the magnitude or even the speed of the spread of this or any other virus.

Furthermore, unless similar testing efforts are made in different countries, the data are not comparable, and pooling them may provide an even worse picture of the spread of the virus worldwide. COVID-19 related deaths data are reported nearly as much, but there is less focus on them. These data, however, are less prone to sampling error [2,3], unless a large number of COVID-19 related deaths go undetected, misdiagnosed, or unreported [10,11]. Therefore, if they can be used to estimate the number of infections, they may provide a much sharper picture of the spread of the disease.

The Infection Fatality Rate (IFR, sometimes referred to as “Infection Fatality Ratio”) can do just that. It is defined as the percentage of cases infected with a given disease, symptomatic or not, that eventually die [12]. Thus, if we know the IFR in a given population and the number of deaths, we can estimate the number of people that were infected around the same date in which those who eventually died got infected. The difference between the number of deaths and such estimate is an estimate of the number of people who have recovered and are now immune. The drawback is that such estimates provide a picture of the spread of the disease up to the date when people reported death today were initially infected. That is IP+ODP days ago, where IP is the Incubation Period and ODP is the Onset to Death Period; estimated to be 5.2 [13,14] and 17.8 d [12] respectively for COVID-19. However, comparing CCD and the number of cases estimated using number of reported deaths even 23 days ago (IP+ODP) may provide a country-specific estimate of the fraction of infections detected by CCD. Such estimates can be compared and calibrated with the picture that antibody surveys should provide, but see Vogel [4], and may even represent a monitoring alternative for countries with little or no access to antibody testing.

Since IFR is age dependent, its global value depends on the age structure of the infected population. COVID-19 is known to be more lethal in older age classes [12,15,16, 17], thus we can expect that the more biased the age structure of a given population to such classes, the higher its IFR will be. In order to apply this value to other populations, the IFR of each age class must be weighted by the relative proportion of each age class in the new population in order to have a corrected estimate of the population IFR.

In this study, we developed a code in the R Statistical Package [18] with functions to estimate and graph: (1) The IFR of different countries by weighting recently published COVID-19 IFR age specific values [12] by the age structure of each country [19]. (2) The number of people infected up to Infection Date (Idt = IP+ODP). (3) A Detection Index (%DI) on Idt as the percentage of confirmed cases to the estimated number of people infected. (4) The percentage of the country’s population already recovered and immune up to the present. (5) The number of people infected between Idt and the present date using %DI. (6) The number of deaths expected from those infections in the following IP+OPD days. (7) The percentage of the country’s population expected to have recovered and be immune in the following IP+ODP days. We show and discuss estimates up to the date of submission for the 50 countries with the highest values of reported deaths. We provide the R code to obtain daily updated estimates for 178 different countries and territories throughout the development of the COVID-19 pandemic.

## Material and methods

A program *(script)* was developed using the R statistical software v 3.6.3 [18]. A fully commented functional version of this script can be downloaded as supporting information (S1 File). Two types of functions are coded, one to graph estimates in countries selected by the user, and another to produce tables with all estimates for countries selected by the user. Relevant constants involved in estimates can be modified in the form of function parameters as explained in S1. It has been recently suggested [20] that sharing fully detailed code and functionality of modeling and monitoring tools is more important than ever to face the current COVID-19 pandemic. This section describes what the script does and the parameters used as default, beginning with the procedure used to obtain the data.

The database used for analyses is obtained from the European Centre for Disease Prevention and Control (ECDC) web page [21]. The script includes code lines needed to import the daily updated database with confirmed cases and deaths. The ECDC keeps a daily updated database curated from over 500 sources [22]. We offer the most recent ECDC database up to the date of submission as supporting information (S2 File). Other daily updated repositories, such as 2019 Novel Coronavirus COVID-19 (2019-nCoV) Data Repository by Johns Hopkins CSSE [23] or the WHO COVID-19 database [24], did not offer the option to automatically download data into a statistical software package. The ECDC database includes reports since December 31, 2019. In contrast, the WHO database offers data since January 8, 2020 and the Johns Hopkins CSSE database since February 23, 2020. No major differences have been detected amongst the three sources [25], but the ECDC has been shown to have the most consistently published and cleanly maintained data [25].

An Incubation Period (IP) of 5.2 d was taken from Li et al. [13] and Backer et al. [14], very close in value to that of Lauer et al. [26] of 5.1 d. An Onset to Death Period (ODP) of 17.8 d was taken from Verity et al. [12]. The sum of these two values (23 d) is subtracted from the date of each daily report to obtain the Infection date (Idt), the date at which people that died on the day reported presumably became infected.

Age-specific Infection Fatality Rates for COVID-19 were first estimated by Team TNCPERE [15]. At least two more studies [12,16] have suggested adjustments to age-specific IFR values considering potential sampling bias in the original report, or non-uniform infection rates of different age classes. We decided to use the values reported by Verity et al. [12], although other alternatives (S3 File) [15,16] can be used instead as IFR values for each age class can also be specified by the user. Age structure data for 178 different countries and territories were obtained from the UN Population Division [19]. The age structure database (provided as S4 File, the R script requires this file) was curated to match the name and geoId code used by the ECDC database in order to estimate each country’s global IFR. A table with each country’s geoId code is also provided for reference (S5 File). Each country’s global IFR is calculated by weighing each age specific IFR by the proportion of the corresponding age class in the country’s population.

A country’s global IFR and daily reported deaths data are used to estimate the daily number of people that were infected on Idt. The premise supporting this procedure is: for a country with a global IFR of X%, for every X number of deaths reported today, 100-X people became infected on the same day (Idt = IP+ODP days ago) and are now presumably immune. The onset to discharge period for COVID-19 cases has been found to be similar or only slightly larger than ODP [12,27]. Cumulative daily estimates of people infected on Idt are obtained by adding all previous daily estimates up to Idt. A country’s daily Detection Index on any given day up to Idt (%DI) is estimated as the CCD value on that date ×100, divided by the estimate of people infected on the same date.

If we assume that infected individuals that did not die recovered and became immune in 23 (IP+ODP) days, in order to estimate how many individuals presently recovered, we need to estimate how many became infected 23 days ago and subtract the present number of deaths. The percentage of each country’s population now recovered and immune is estimated by subtracting the present cumulative number of deaths from the present cumulative estimate of people infected up to Idt ×100, and dividing by the reported country’s population in 2018 (data form The World Bank Group included in the ECDC database [21]). This value estimates the present percentage of the country’s population represented by the estimate of the number of people that became infected, but survived, up to the date in which the deaths reported today became infected.

Since the estimated number of infections depends on the number of deaths projected IP+ODP days back, the previous procedure does not allow for estimating the number of people infected between Idt and the present. In order to attempt an estimate of infections up to the present, CCD and %DI are used assuming %DI remains constant from Idt to the present under this premise: for a country with %DI of Y%, for every Y confirmed cases, there are 100-Y more undetected infections. Given that daily %DI varies between days and over time throughout the pandemic (see below), and in order to have the most recent %DI estimate, the %DI value reported and used to estimate the daily number of infections from Idt to the present is the percentage of total estimated infections in a seven day period ending with Idt represented by total CCD in the same period. This value will be referred to as %DI on Idt. If a country is currently reporting no deaths, this %DI value is evaluated for the period ending with the last daily %DI value available. The script also estimates infections from Idt to the present under two scenarios: a two-fold increase of %DI on Idt (%DI×2), and a decrease of 50% (%DI/2). Daily estimates are added to cumulative estimates up to Idt to obtain present cumulative estimates.

Daily infection estimates from Idt to the present are multiplied by IFR in order to obtain daily estimates of death and survivors from the present up to IP+ODP days in the future. The daily estimated number of deaths is added to the present number of deaths reported in order to estimate the future cumulative number of deaths expected from the present up to IP+ODP days in the future. Likewise, the daily estimated number of survivors is added to the number of survivors estimated up to the present to estimate the future percentage of recovered and immune population in the same period. The same scenarios of increase and decrease in the country’s %DI after Idt are considered to provide estimation ranges for the future cumulative number of deaths and for the future percentage of recovered and immune population. Finally, the script calculates the increase percentage by which cumulative number of estimated infections have increased from Idt to the present, the same percentage by which future deaths and percentage of immune population are estimated to increase in 23 days.

We obtained estimates for the 50 countries with the greatest number of accumulated deaths on the date of final submission, representing 98.53% of the total number of deaths worldwide (Table 1 and S6 File), and figures representing the trajectory of these estimates in four representative countries: Belgium, United States, Brazil and Japan (these and all the remaining 46 country graphs can be found in the supporting information as S7 File).

**Table 1.**
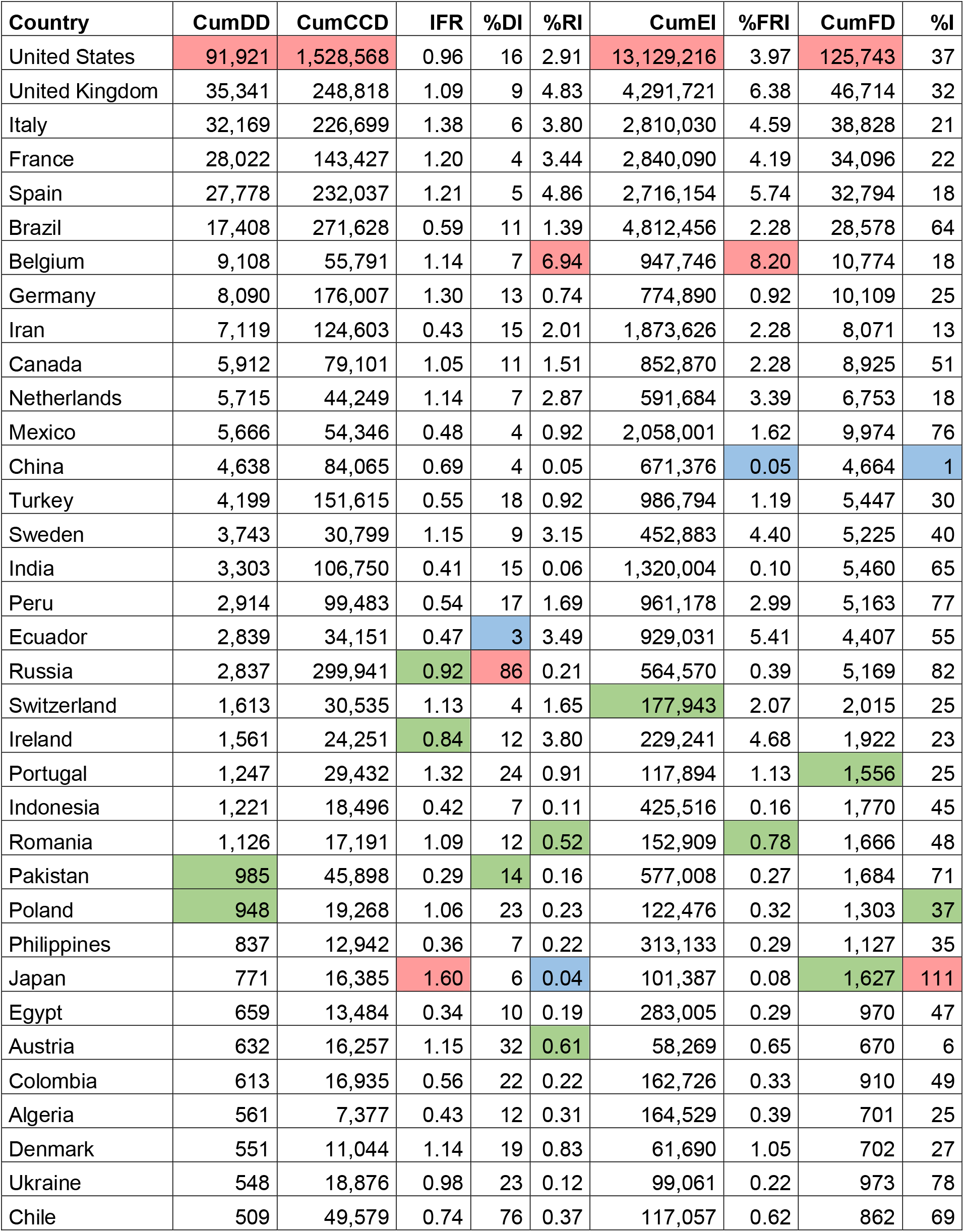
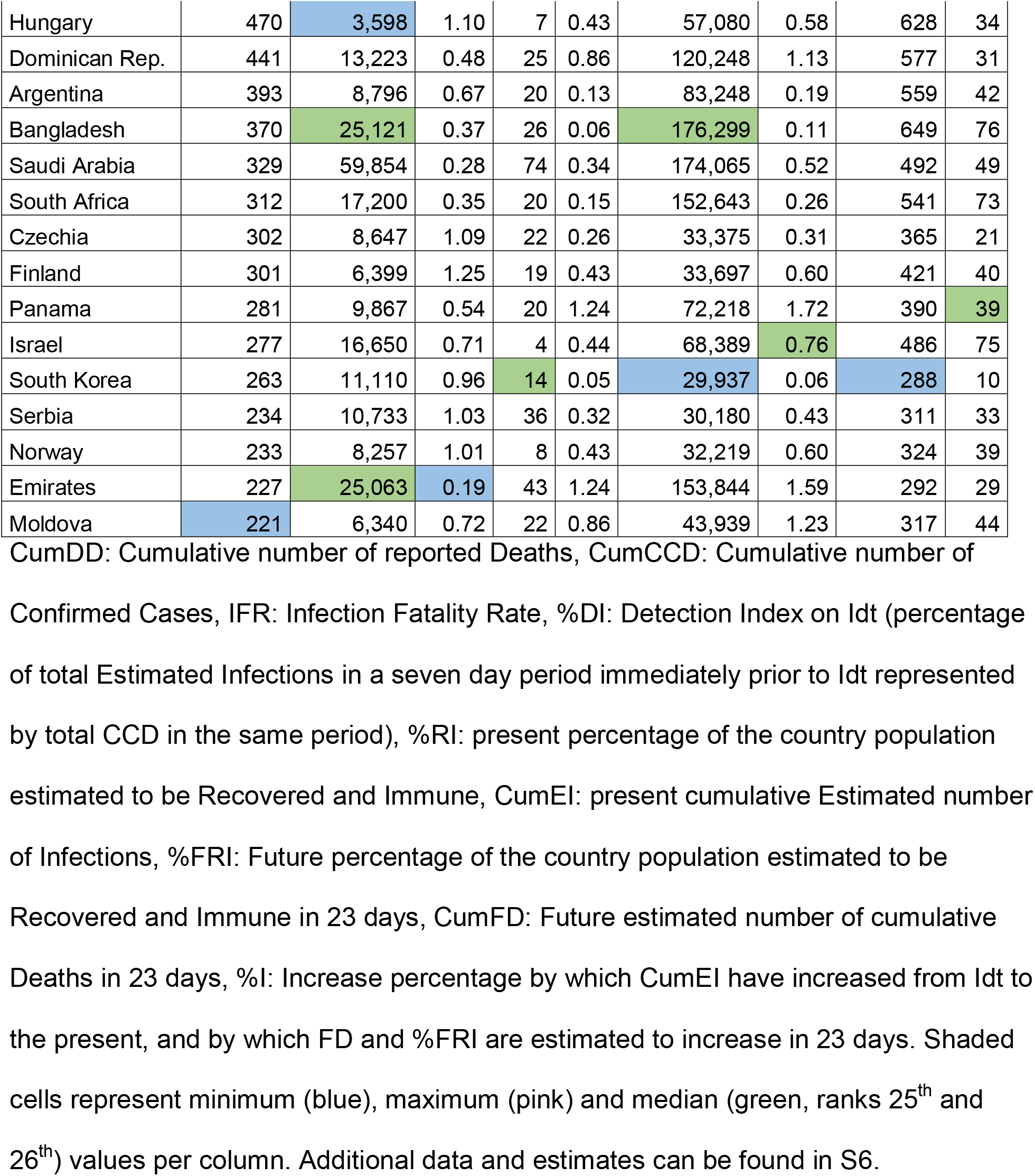
Data and estimates for the 50 countries with the greatest number of accumulated deaths during the COVID-19 pandemic on May 20, 2020.

| Country | CumDD | CumCCD | IFR | %DI | %RI | CumEI | %FRI | CumFD | %I |
| --- | --- | --- | --- | --- | --- | --- | --- | --- | --- |
| United States | 91,921 | 1,528,568 | 0.96 | 16 | 2.91 | 13,129,216 | 3.97 | 125,743 | 37 |
| United Kingdom | 35,341 | 248,818 | 1.09 | 9 | 4.83 | 4,291,721 | 6.38 | 46,714 | 32 |
| Italy | 32,169 | 226,699 | 1.38 | 6 | 3.80 | 2,810,030 | 4.59 | 38,828 | 21 |
| France | 28,022 | 143,427 | 1.20 | 4 | 3.44 | 2,840,090 | 4.19 | 34,096 | 22 |
| Spain | 27,778 | 232,037 | 1.21 | 5 | 4.86 | 2,716,154 | 5.74 | 32,794 | 18 |
| Brazil | 17,408 | 271,628 | 0.59 | 11 | 1.39 | 4,812,456 | 2.28 | 28,578 | 64 |
| Belgium | 9,108 | 55,791 | 1.14 | 7 | 6.94 | 947,746 | 8.20 | 10,774 | 18 |
| Germany | 8,090 | 176,007 | 1.30 | 13 | 0.74 | 774,890 | 0.92 | 10,109 | 25 |
| Iran | 7,119 | 124,603 | 0.43 | 15 | 2.01 | 1,873,626 | 2.28 | 8,071 | 13 |
| Canada | 5,912 | 79,101 | 1.05 | 11 | 1.51 | 852,870 | 2.28 | 8,925 | 51 |
| Netherlands | 5,715 | 44,249 | 1.14 | 7 | 2.87 | 591,684 | 3.39 | 6,753 | 18 |
| Mexico | 5,666 | 54,346 | 0.48 | 4 | 0.92 | 2,058,001 | 1.62 | 9,974 | 76 |
| China | 4,638 | 84,065 | 0.69 | 4 | 0.05 | 671,376 | 0.05 | 4,664 | 1 |
| Turkey | 4,199 | 151,615 | 0.55 | 18 | 0.92 | 986,794 | 1.19 | 5,447 | 30 |
| Sweden | 3,743 | 30,799 | 1.15 | 9 | 3.15 | 452,883 | 4.40 | 5,225 | 40 |
| India | 3,303 | 106,750 | 0.41 | 15 | 0.06 | 1,320,004 | 0.10 | 5,460 | 65 |
| Peru | 2,914 | 99,483 | 0.54 | 17 | 1.69 | 961,178 | 2.99 | 5,163 | 77 |
| Ecuador | 2,839 | 34,151 | 0.47 | 3 | 3.49 | 929,031 | 5.41 | 4,407 | 55 |
| Russia | 2,837 | 299,941 | 0.92 | 86 | 0.21 | 564,570 | 0.39 | 5,169 | 82 |
| Switzerland | 1,613 | 30,535 | 1.13 | 4 | 1.65 | 177,943 | 2.07 | 2,015 | 25 |
| Ireland | 1,561 | 24,251 | 0.84 | 12 | 3.80 | 229,241 | 4.68 | 1,922 | 23 |
| Portugal | 1,247 | 29,432 | 1.32 | 24 | 0.91 | 117,894 | 1.13 | 1,556 | 25 |
| Indonesia | 1,221 | 18,496 | 0.42 | 7 | 0.11 | 425,516 | 0.16 | 1,770 | 45 |
| Romania | 1,126 | 17,191 | 1.09 | 12 | 0.52 | 152,909 | 0.78 | 1,666 | 48 |
| Pakistan | 985 | 45,898 | 0.29 | 14 | 0.16 | 577,008 | 0.27 | 1,684 | 71 |
| Poland | 948 | 19,268 | 1.06 | 23 | 0.23 | 122,476 | 0.32 | 1,303 | 37 |
| Philippines | 837 | 12,942 | 0.36 | 7 | 0.22 | 313,133 | 0.29 | 1,127 | 35 |
| Japan | 771 | 16,385 | 1.60 | 6 | 0.04 | 101,387 | 0.08 | 1,627 | 111 |
| Egypt | 659 | 13,484 | 0.34 | 10 | 0.19 | 283,005 | 0.29 | 970 | 47 |
| Austria | 632 | 16,257 | 1.15 | 32 | 0.61 | 58,269 | 0.65 | 670 | 6 |
| Colombia | 613 | 16,935 | 0.56 | 22 | 0.22 | 162,726 | 0.33 | 910 | 49 |
| Algeria | 561 | 7,377 | 0.43 | 12 | 0.31 | 164,529 | 0.39 | 701 | 25 |
| Denmark | 551 | 11,044 | 1.14 | 19 | 0.83 | 61,690 | 1.05 | 702 | 27 |
| Ukraine | 548 | 18,876 | 0.98 | 23 | 0.12 | 99,061 | 0.22 | 973 | 78 |
| Chile | 509 | 49,579 | 0.74 | 76 | 0.37 | 117,057 | 0.62 | 862 | 69 |
| Hungary | 470 | 3,598 | 1.10 | 7 | 0.43 | 57,080 | 0.58 | 628 | 34 |
| Dominican Rep. | 441 | 13,223 | 0.48 | 25 | 0.86 | 120,248 | 1.13 | 577 | 31 |
| Argentina | 393 | 8,796 | 0.67 | 20 | 0.13 | 83,248 | 0.19 | 559 | 42 |
| Bangladesh | 370 | 25,121 | 0.37 | 26 | 0.06 | 176,299 | 0.11 | 649 | 76 |
| Saudi Arabia | 329 | 59,854 | 0.28 | 74 | 0.34 | 174,065 | 0.52 | 492 | 49 |
| South Africa | 312 | 17,200 | 0.35 | 20 | 0.15 | 152,643 | 0.26 | 541 | 73 |
| Czechia | 302 | 8,647 | 1.09 | 22 | 0.26 | 33,375 | 0.31 | 365 | 21 |
| Finland | 301 | 6,399 | 1.25 | 19 | 0.43 | 33,697 | 0.60 | 421 | 40 |
| Panama | 281 | 9,867 | 0.54 | 20 | 1.24 | 72,218 | 1.72 | 390 | 39 |
| Israel | 277 | 16,650 | 0.71 | 4 | 0.44 | 68,389 | 0.76 | 486 | 75 |
| South Korea | 263 | 11,110 | 0.96 | 14 | 0.05 | 29,937 | 0.06 | 288 | 10 |
| Serbia | 234 | 10,733 | 1.03 | 36 | 0.32 | 30,180 | 0.43 | 311 | 33 |
| Norway | 233 | 8,257 | 1.01 | 8 | 0.43 | 32,219 | 0.60 | 324 | 39 |
| Emirates | 227 | 25,063 | 0.19 | 43 | 1.24 | 153,844 | 1.59 | 292 | 29 |
| Moldova | 221 | 6,340 | 0.72 | 22 | 0.86 | 43,939 | 1.23 | 317 | 44 |

CumDD: Cumulative number of reported Deaths, CumCCD: Cumulative number of Confirmed Cases, IFR: Infection Fatality Rate, %DI: Detection Index on Idt (percentage of total Estimated Infections in a seven day period immediately prior to Idt represented by total CCD in the same period), %RI: present percentage of the country population estimated to be Recovered and Immune, CumEI: present cumulative Estimated number of Infections, %FRI: Future percentage of the country population estimated to be Recovered and Immune in 23 days, CumFD: Future estimated number of cumulative Deaths in 23 days, %I: Increase percentage by which CumEI have increased from Idt to the present, and by which FD and %FRI are estimated to increase in 23 days. Shaded cells represent minimum (blue), maximum (pink) and median (green, ranks 25<sup>th</sup> and 26<sup>th</sup>) values per column. Additional data and estimates can be found in S6.

## Results and discussion

Estimated IFR values show a bimodal distribution with one mode around 0.5% and the other slightly above 1% (Fig 1). Out of the 50 countries with more reported deaths, estimated IFR values are higher than 1% in all European countries (n=23) with the exception of Russia (0.92%), Ireland (0.84%), Ukraine (0.98%) and Moldova (0.72%), and lower than 1% in all non-European countries (n=27) with the exception of Canada (1.05%) and Japan (1.60%) (Table 1). This is due to the fact that European countries have age structures biased to higher age classes. This is likely one of the reasons why European countries have had high death tolls, and why mortality may be lower in other continents [17, 28], although many other additional reasons have been suggested [29-31].

**Fig 1.**
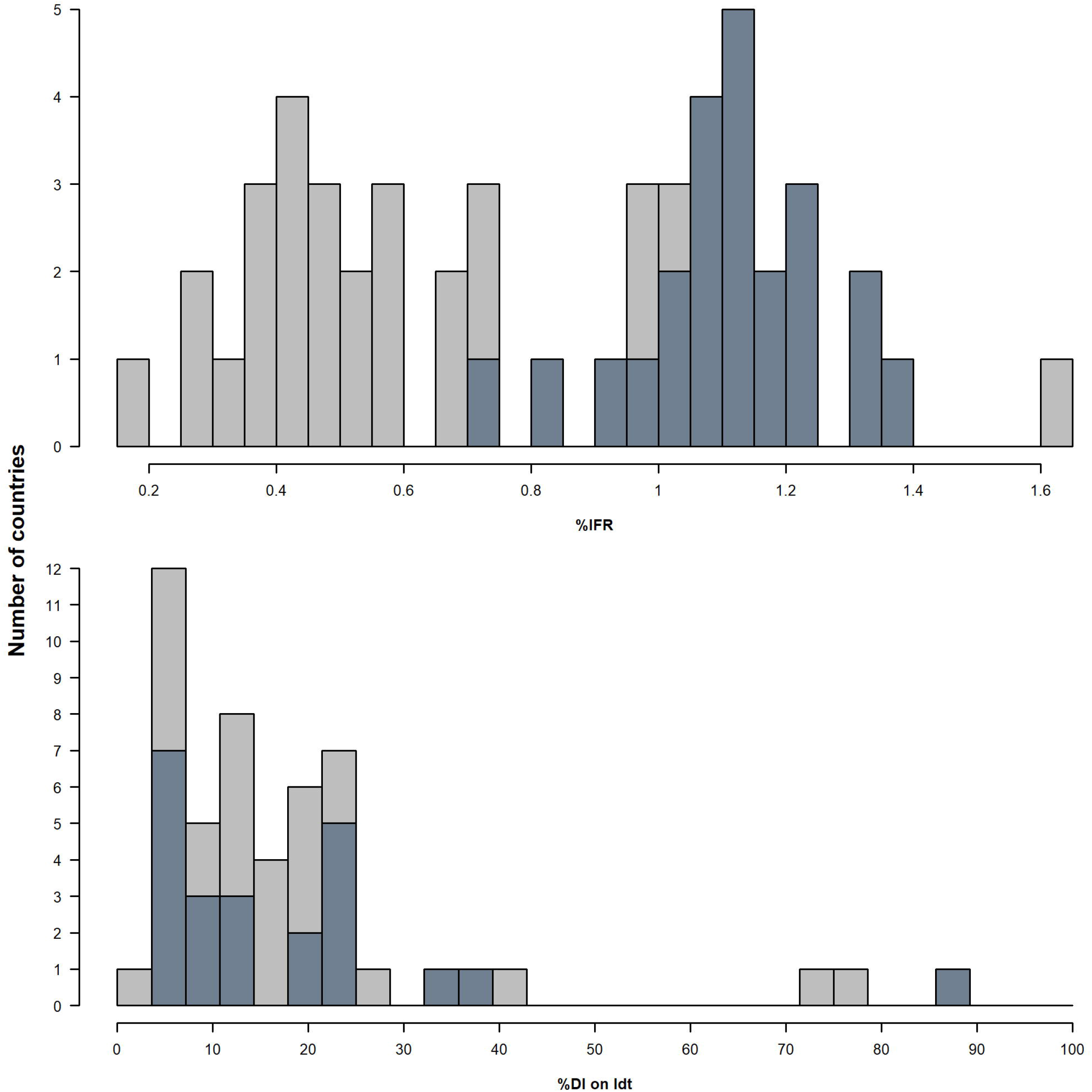
Distribution of IFR and %DI estimates in the 50 countries with more reported deaths during the development of the COVID-19 pandemic up to May 20, 2020. Upper panel: Infection Fatality Rate (IFR) seems to be bimodal, with most European countries (darker bars) showing values above 1%, and most of the remaining countries with values below 1%. Lower panel: In most countries the percentage of total Estimated Infections in a seven day period immediately prior to Infection Date represented by total confirmed cases in the same period (%DI on Idt) is under 30%. European countries highlighted as darker bars.

Our estimates up to Idt (estimated infections, %DI on Idt and percentage of immune population) depend only on the number of reported deaths, age specific IFR, age structure data and population size, so they are as good as these data values are. Assuming age structure and population size data are reliable and that the number of COVID-19 related deaths are as accurate as they should be [2,3,11], age specific IFR values remain a key element of our estimations. Verity et al [12] have, in our opinion, made every possible effort to correct for sampling bias in the Crude Case Fatality Rate from Wuhan data [15] in order to obtain true IFR values for each 10-year age class; therefore we have used such values to estimate global IFR for each country. However, IFR may also depend on country specific factors such as healthcare system, socio-economic structure and/or proportion and types of comorbidities [29,30].

This paper will not address possible reasons why different countries have shown different trajectories in terms of CCD and reported deaths other than differences in IFR; instead we attempt to identify other useful estimates and patterns common to several countries, and their implications. The results discussed in this manuscript will have changed somewhat within days as CCD and reported deaths data are updated every day, but the patterns are likely to remain, as we have observed throughout the previous weeks working on this manuscript. However, Table 1, S6 File and all figures may be updated every day with the functions in the provided R script. We report estimates on absolute numbers of past and present infections, and future deaths, as they may be useful for individual countries. However, we focus on discussing relative values derived from them (IFR, %DI, present and future estimates of immune population and its predicted percentage increase) as they can be compared across countries regardless of their size.

Our results on %DI on Idt show great variation among countries, and suggest that CCD represent less than a third of estimated infections on Idt in all but 5 countries, Russia (86%), Chile (76%), Saudi Arabia (74%) United Arab Emirates (43%) and Serbia (36%) (Fig 1 and Table 1). Very high values of %DI (either daily values or on Idt), even above 100%, may be obtained occasionally for reasons other than high testing effort. If not all deaths are detected or reported [10], our estimates based on reported deaths and IFR may underestimate the number of infections, thus overestimating %DI. Another possibility is that one or more age specific IFR values are lower in certain countries from those suggested by Verity et al. [12]. Since IP and ODP are average values with important variation [12,13], the number of reported deaths on a given day must certainly include infections on other dates around Idt. Nevertheless, we recommend that original data should be verified, and age specific IFR values be evaluated, in countries with %DI values consistently far outside the distribution of the rest of the countries (Fig 1). Other potential outliers in the estimates may be due to occasional corrections in the ECDC database, such as a change of methodology taken by Ecuador’s healthcare system on May 8 [32], which resulted in negative CCD values.

In spite of variation in daily %DI values, a clear pattern of decrease throughout the pandemic may be observed among most of the 50 countries (orange points in Figs 2-4 and S7), suggesting that testing effort is not, or cannot be, maintained by most countries once the curve has taken off. A few countries show long-term fluctuations in daily %DI values such as Japan (Fig 5), having reached daily %DI values near 100% in early April, but with a %DI on Idt of 6%. Although correction factors have been used to estimate actual number of infections from CCD [33], the patterns of short and long term fluctuation observed suggest that CCD do not provide a consistent estimate of the actual number of infections at any point in time.

**Fig 2.**
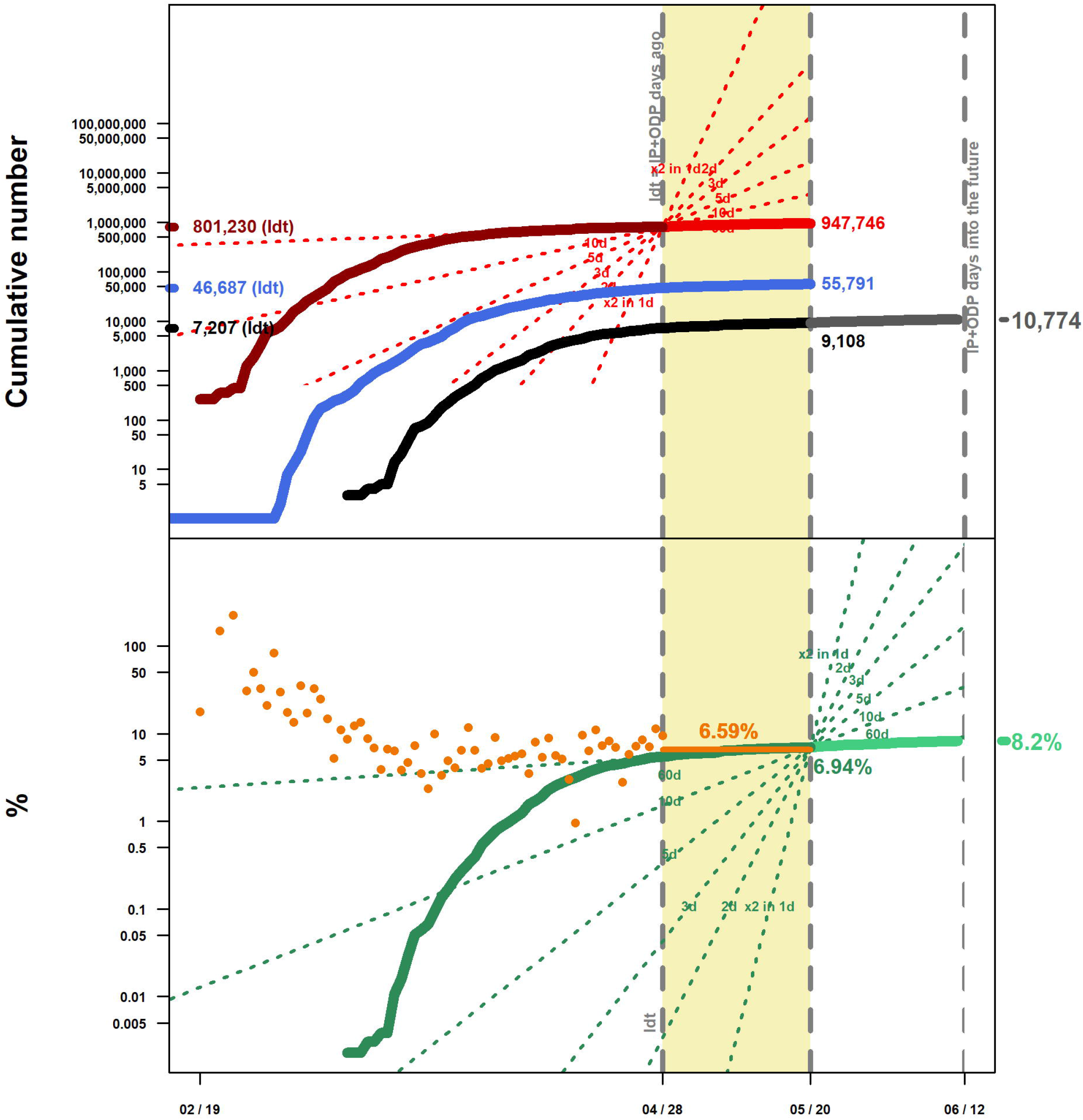
Data and estimates during the development of the COVID-19 pandemic in Belgium up to May 20, 2020. Upper panel: Cumulative number of reported deaths (black line) and confirmed cases (blue line) up to the present, according to the European Centre for Disease Prevention and Control. Estimated infections up to Infection Date (Idt, April 28, dark red line). Estimated infections from Idt up to the present (light red line), assuming a constant Detection Index after Idt. Estimated cumulative number of deaths up to June 12 (grey line), assuming a constant Detection Index after Idt. Lower panel: Daily Detection Index (orange points) expressing the daily percentage of estimated infections represented by the daily number of Confirmed Cases, and its estimated value during the period of seven days prior to Idt (orange horizontal line). Percentage of the country population estimated to have recovered and be immune up to the present (dark green line), and up to June 12 assuming a constant Detection Index after Idt (light green line). Values in the corresponding colors represent totals up to either Idt (on Y axis in upper panel) or up to the ending date of each line. Dotted convergent lines show slopes for theoretical two-fold increases in 1, 2, 3, 5, 10 and 60 days. Vertical grey broken lines represent Idt (April 28), present (May 20) and IP (Incubation Period) + ODP (Onset to Death Period) days into the future (June 12). Belgium has experienced high relative mortality, a low detection index, and the highest value of estimated percentage of recovered and immune population. After a very steep beginning, the rate of increase of all estimates is now very low. These parameters are expected from countries in which, after the disease has become widespread, the rate of infections has decreased substantially.

**Fig 3.**
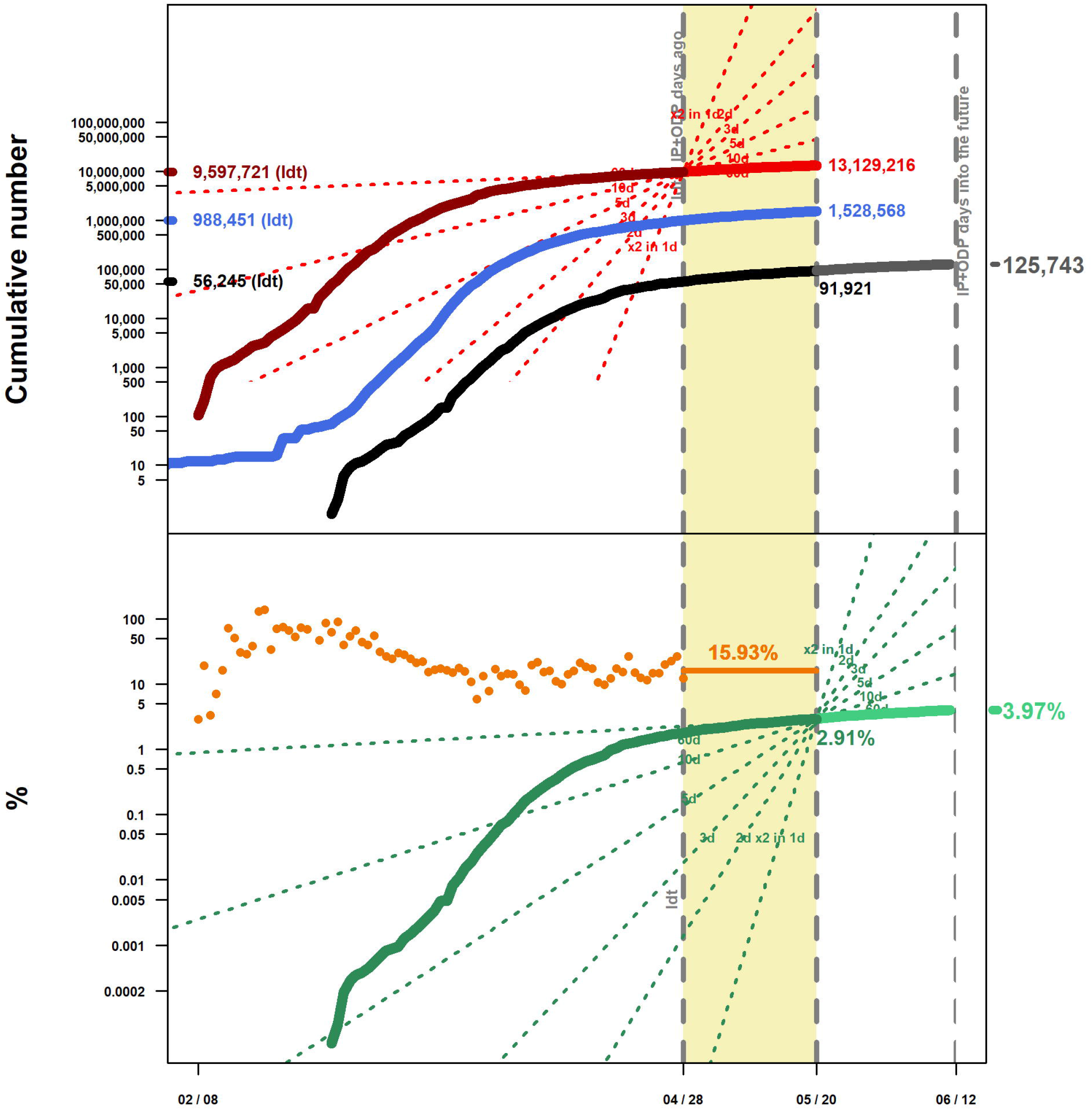
Data and estimates during the development of the COVID-19 pandemic in the United States up to May 20, 2020. For abbreviation meanings and code color explaining each line see legend in Fig 2. The United States has the highest COVID-19 death toll of the world, a still clear ascending pattern, moderate detection index and also a still moderate estimated percentage of immune population.

**Fig 4.**
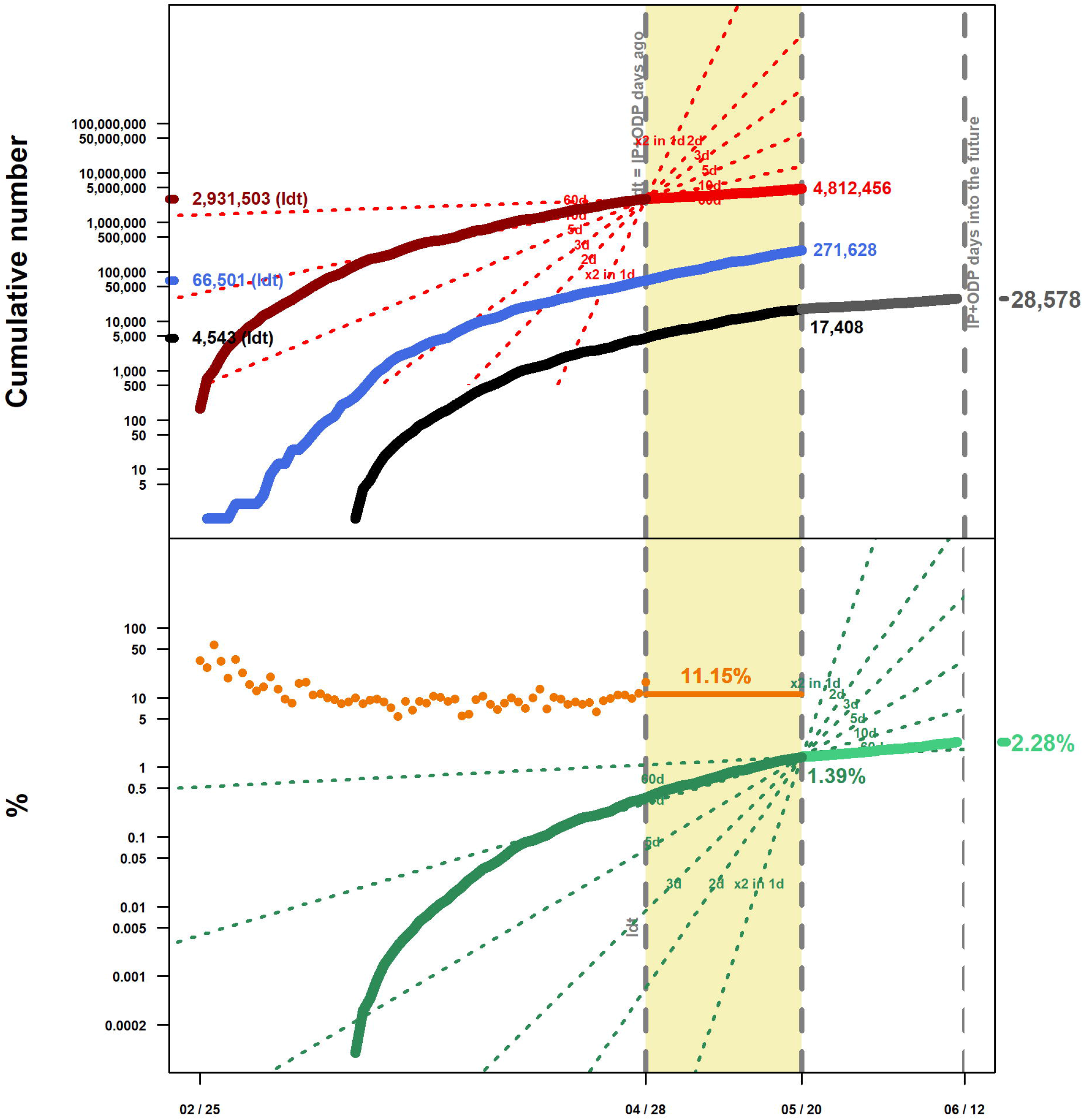
Data and estimates during the development of the COVID-19 pandemic in Brazil up to May 20, 2020. For abbreviation meanings and code color explaining each line see legend in Fig 2. With an already high death toll and a low Detection Index, estimates for Brazil are still increasing at relatively high rates. These parameters are expected from countries still facing a steep increase in infections, death toll and population immunity.

**Fig 5.**
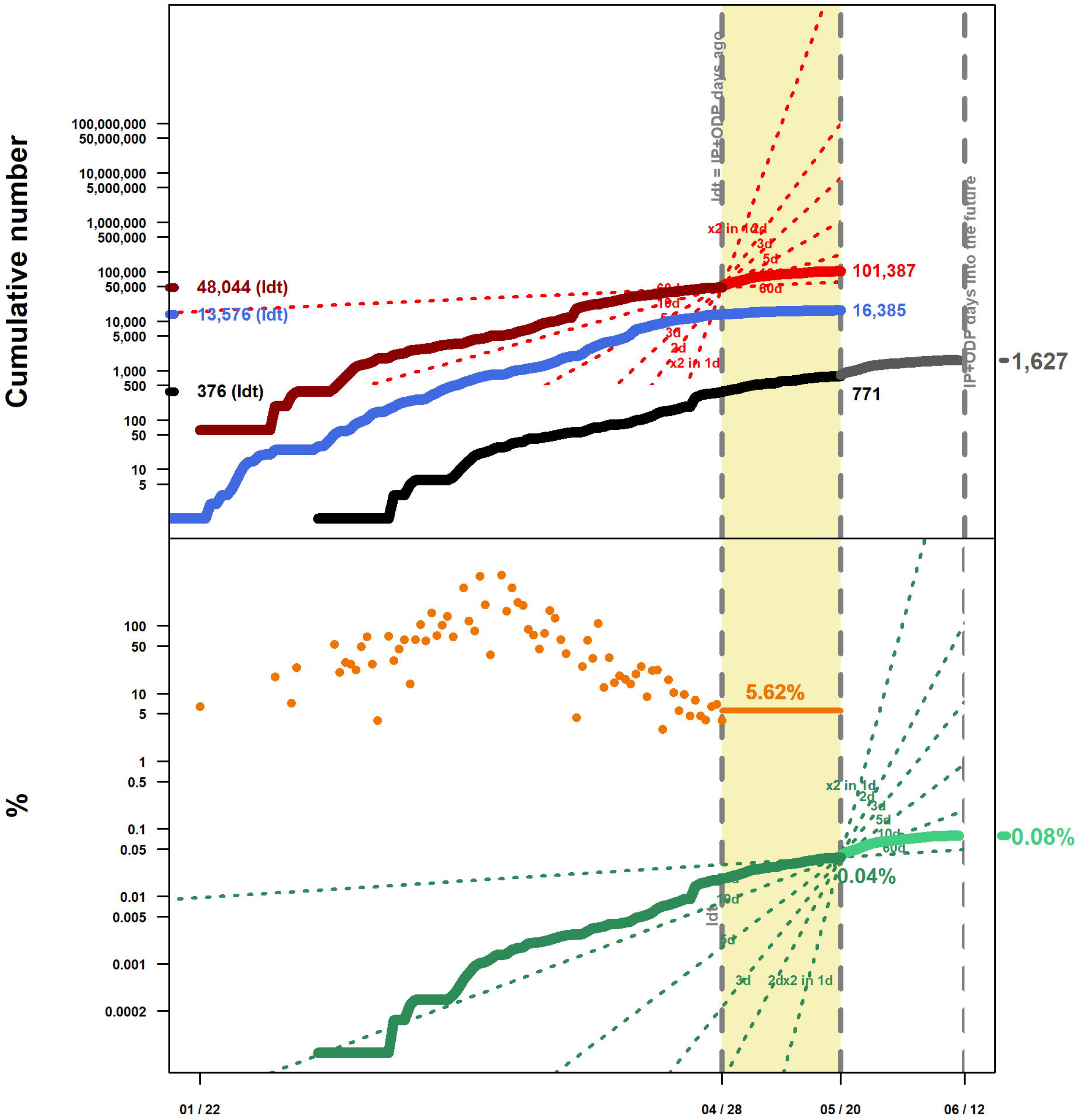
Data and estimates during the development of the COVID-19 pandemic in Japan up to May 20, 2020. For abbreviation meanings and code color explaining each line see legend in Fig 2. With the lowest percentage of immune population, Japan has experienced long term fluctuation in daily %DI values throughout the pandemic but its %DI value on Idt is low. The flat curves for data and estimates throughout most of the pandemic and the low value of estimated percentage of immune population are expected from countries that have kept the spread under control from the beginning.

CCD are derived from SARS-CoV-2 PCR specific tests [3,34,35] performed seeking several goals. Probably the main goal is, or should be, to identify currently infected people (i.e. carrying the virus either with symptoms or not) in order to track and contain other infections [3,35]. As these results accumulate, and given that some countries carry out tests in a more or less standardized fashion, it is tempting to report them (as many countries do) as a measure of the spread of the SARS-CoV-2 virus either directly, or as the relative number of positive outcomes. However, there are several reasons why this is not appropriate. They are rarely applied to a random sample of the population, but rather aimed at suspicious individuals (travelers or recent contacts with other infections) or at symptomatic individuals [6, 12, 27]. These tests will have a negative outcome if the virus was present, but is not anymore [6, 9]. To solve these issues, there are current efforts to develop and apply lateral flow immunoassays to measure the seroprevalence of antibodies to SARS-CoV-2 to random samples of the population, such as the work carried out by Bendavid et al [6]. This study suggests a 50-85 fold increase in the number of infections compared to CCD, but see Vogel [4]. Comparing our estimates with antibody surveys once they are commonly implemented should provide the ability to calibrate them, adjusting country and/or age specific IFR values. A recent antibody survey in Spain [36] reported an immunity percentage of people tested between April 27 and May 11 of 5% (95% IC: 4.7-5.4%), in close agreement with our current estimate of 4.86% for this country (Table 1).

A strong investment by a given country in PCR-based analyses may be seen as the country’s commitment to identify and contain lines of infection, and thus control the spread of the virus. A byproduct of a high testing effort would be a high %DI value. However, a high %DI value may be hard to reach or maintain for any country once the virus has spread in large numbers. Most (17) of the top 25 countries in Table 1, those with the highest death tolls and thus the highest estimated number of infections, have %DI values on Idt under the median (14%), while most (18) of the remaining countries, those with much lower values of accumulated deaths and thus lower estimated number of infections, have values on Idt above the median.

The six countries with estimated current percentages of population recovered and immune above 3% and IFR above 1% (United Kingdom, Italy, France, Spain, Belgium Fig 2 and Sweden), are those well known for their initially steep slope of increase in CCD and in number of reported deaths, and the resulting high death toll. The United States (Fig 3) and South Korea, having both a global IFR of 0.96, have clearly contrasting estimated percentages of immune population (2.91 *vs* 0.05%). This suggests that percentages of immune population in countries where the lines of infection could not be controlled will grow more rapidly and may reach up to 8.2% in the next 23 days (Belgium, Fig 2 and Table 1). Our results suggest that other countries are still experiencing a steeper increase in number of infections (see Increase percentage column in Table 1). A group of five countries (Brazil Fig 4, México, India, Peru and Russia) with IFR values below 1% and an already high death toll (above 1,000) may experience a high number of casualties if, as our estimates suggest, they experience 68-82% more deaths in the next 23 days. They would also experience an equivalent increase in population immunity.

It has been suggested that the accumulation of herd immunity in the population slows epidemic resurgence [37]. By contrast, when virtually no population immunity is built, such as the case of China, India, Japan, Bangladesh or South Korea (percentage of immune population below 0.1%, in Table 1) a resurgence peak of COVID-19 may be nearly the same size as an uncontrolled epidemic if control policies fail [37]. The numbers for Japan (Fig 5), for instance, suggest a high vulnerability since it shows the highest estimated IFR (1.60) of all countries, a low %DI on Idt (6 %), the lowest percentage of immune population (0.04%), and the highest estimated increase (111%) in infections from Idt to the present, and thus a similar increase in deaths in the following 23 days. We still don’t know how long immunity to SARS-CoV-2 could last [4], but recent modeling suggests anywhere between 40 weeks and 5 years [37]. Nevertheless, immunity is unlikely to end abruptly, so the rate of any further re-infection with new strains should be smoother in the future as long as *herd immunity* has had some build-up [4,16]. However, even Belgium with the highest estimated percentage of immune population (6.94%) seems far from values approaching herd immunity.

Although CCD have been used to estimate the number of infections [38], we believe that this will usually provide poor estimates as suggested by the short and long-term variation in daily %DI values. In any case, we suggest that having a low %DI value means that a large proportion of infections is not being tracked. Other values that are calculated based on CCD are likely just as misleading, such as Case Fatality Rate [2,3,6] and Recovered Cases. A key factor for the gap between CCD and the real number of infections are asymptomatic infections (which technically do not suffer COVID-19 so they are not “cases”, but asymptomatic active vectors of SARS-CoV-2) [7,8]. In the absence of widespread and well-designed application of lateral flow immunoassays to random samples of the population to measure the seroprevalence of antibodies, we suggest that estimates based on reported deaths and IFR are a more reliable alternative to estimate the spread of SARS-CoV-2 than CCD in any country for which age structure data is available and data of reported deaths is trustworthy. They also illustrate the potential bias when assuming that CCD data reflect the actual spread of COVID-19.

## Data Availability

All data used are included as supplementary files.

## Acknowledgements

We are grateful to Isabel Noriega, Yareni Perroni, Matt Draud and Warren Greiff for proofreading and useful comments on early versions of the manuscript.

## Supporting information

**S1 File. Fully explained R Script** (.R)

**S2 File. COVID-19 geographic distribution worldwide on May 20, 2020**. Dataset downloaded from the ECDC website. (.XLSX)

**S3 File. IFR by age from three sources** (.DOCX)

**S4 File. Age structure of 178 countries**. The estimated 2020 country age structure by the UN Population Division, curated to match the geoId code used by the ECDC database. (.TXT)

**S5 File. GeoId codes**. For reference, geoId codes and full country names, obtained from the ECDC website. (.CSV)

**S6 File. Extended results table of data and estimates during the development of the COVID-19 pandemic up to May 20, 2020, for the 50 countries with more reported deaths**. This spreadsheet is generated by the function cv19.tab.num() in the R script (S1) and includes 10 additional columns apart from those shown in Table 1: (1) the cumulative number of Confirmed Cases on Infection date (CCD-Idt), (2) the Estimated number Infections on Infection date (EI-Idt), (3) EI assuming a two-fold %DI increase (EI %DIx2), and (4) assuming a %DI decrease of 50% (EI %DI/2), (5-6) future recovered and immune infections (%FRI %DIx2; %FRI %DI/2) and (7-8) future cumulative number of deaths (FD %DIx2; FD %DI/2) under the same two scenarios of %DI increase and decrease, and (9-10) days since 5 deaths (d5d) and since 100 deaths (d100d) were reported for each country (.CSV). Relevant parameters used by the functions are shown at the bottom of the table.

**S7 File. Graphs of data and estimates during the development of the COVID-19 pandemic up to May 20, 2020, for the 50 countries with more reported deaths**. This PDF file is generated by the function cv19.plot.num() in the R script (S1). A list of countries included, their geoId codes and relevant parameters used are shown in the first page.

